# A DSM-5 AMPD and ICD-11 compatible measure for an early identification of personality disorders in adolescence – LoPF-Q 12-18 latent structure and short form

**DOI:** 10.1101/2022.05.19.22275330

**Authors:** Ronan Zimmermann, Martin Steppan, Johannes Zimmermann, Lara Oeltjen, Marc Birkhölzer, Klaus Schmeck, Kirstin Goth

## Abstract

The LoPF-Q 12-18 (Levels of Personality Functioning Questionnaire) was designed for clinical use and to promote early detection of personality disorder (PD). It is a self-report measure with 97 items to assess personality functioning in adolescents from 12 years up. It operationalizes the dimensional concept of personality disorder (PD) severity used in the Alternative DSM-5 Model for Personality Disorders and the ICD-11. In this study, we investigated the factorial structure of the LoPF-Q 12-18. Additionally, a short version was developed to meet the need of efficient screening for PD in clinical and research applications.

To investigate the factorial structure, several confirmatory factor analysis models were compared. A bifactor model with a strong general factor and four specific factors showed the best nominal fit (CFI = .91, RMSEA = .04, SRMR = .07).

The short version was derived using the ant colony optimization algorithm. This procedure resulted in a 20-item version with excellent fit for a hierarchical model with four first order factors to represent the domains and a secondary higher order factor to represent personality functioning (CFI = .98, RMSEA = .05, SRMR = .04). Clinical validity (effect size d= 3.1 between PD patients and controls) and clinical utility (cutoff ≥ 36 providing 87.5% specificity and 80.2% sensitivity) for detecting patients with PD were high for the short version. Both, the long and short LoPF-Q 12-18 version are ready to be used for research and diagnostic purposes.

## Introduction

### Recent changes in the conceptualisation of personality disorders (PD)

The conceptualisation of personality disorder (PD) and, subsequently, the diagnostic system for PDs is currently transitioning from a categorical system (e.g. narcissistic or avoidant PD) which is still the official system in the DSM-5 [1] to a dimensional approach. The dimensional approach is used both in the ICD-11 [2] as well as in the Alternative DSM-5 Model for Personality Disorders (AMPD). The AMPD is described in the section “emerging measures and models” of the DSM-5. The reason for the fundamental change of the guidelines for diagnosing PD are well documented shortcomings of the categorical system: For instance, individual differences in the expression of PD characteristics are not dichotomous but appear continuously distributed, thresholds (i.e. number of present symptoms required to assign a diagnosis) for categorical PD diagnoses have been criticised as largely arbitrary, and the empirical covariation of the individual criteria does not fully correspond with their assignment to the ten distinct PD categories in the diagnostic manual [3]. As a consequence, the categorical approach is no longer regarded as the only valid taxonomy and has been criticised as a hindrance to research and practice [4]. A growing number of publications in the field now argue in favour of the dimensional approach.

The AMPD and the ICD-11 dimensional PD models are conceptually similar. They each contain two assessment modules to characterise PDs: A first diagnostic module is the evaluation of the self- and the interpersonal functioning of the patients to represent general features and the severity of the PD. This is referred to as ‘criterion A’ in the AMDP. Criterion A is constructed from four domains: identity, self-direction (those account for self-related functioning), empathy and intimacy (accounting for interpersonal functioning). The second module is used to evaluate maladaptive personality traits to represent stylistic differences in the expression of PD [5]. This assessment of ‘trait domain qualifiers’ in the ICD-11 is not mandatory for a diagnosis but helps to refine the assessment. In contrast, in the AMPD, the usage of this second module, ‘Criterion B’, is required.

As a further major shift in the PD diagnosis paradigm, experts now broadly agree on the importance of early detection and treatment of PD [6]. Strong evidence has been delivered showing that PD is a valid diagnosis in youth [7]. PDs can have their origin during childhood and can emerge in early adolescence [8,9]. Early detection and treatment are important as adolescence is a critical and formative period which lays the foundation in terms of psychosocial functioning for the wellbeing and productivity of the adult [10]. Additionally, from a neurocognitive perspective, adolescence also represents a window of opportunity to effectively and efficiently treat mental disorders [8]. By providing early interventions in adolescents, clinicians are trying to avert the harmful psychosocial consequences of a developing disorder and prevent chronification. This is important as PDs can have a heavily incapacitating impact on the patients and their environment, including the somatic health and life expectancy of those affected [11].

Additionally, societal costs of untreated PD are high (e.g. direct healthcare costs and loss of productivity) [12,13]. To account for a perspective of PD across lifetime, both the ICD-11 and the AMPD have abolished the age limit for PD diagnoses. Multiple manualised psychotherapies are available for young PD patients [14–17]. However, to allow for early treatment, age-adequate assessment procedures to detect PDs in adolescents according to the dimensional PD concept are required.

### Need for psychometric instruments

These two-fold changes (dimensional approach and earlier diagnosis) in the diagnostic systems of PDs pose a challenge for mental health care services on a global level. The World Health Organization emphasises the ICD-11 system for PDs needs to be useful and usable also for health care workers in lower-resource settings who are not highly trained specialists [18]. To overcome this challenge, evidence-based assessments are of critical importance (19). Zimmermann et al. [5] provide a brief but comprehensive review on research regarding the dimensional PD models summarizing currently available measures. Birkhölzer et al. [19] provide an updated review for instruments to measure criterion A.

The ICD-11 model for PD is relatively new. Tools specifically targeting the ICD-11 operationalisation of PD diagnosing are currently being developed and validated. Based on strong similarities of ICD-11 and the AMPD regarding personality functioning, Bach & First [20] propose that assessment tools developed for the DSM-5 AMPD model can also be used to support an ICD-11 dimensional PD diagnosis. To comply with the new ICD-11 lifetime perspective on mental disorders in general, all psychometric instruments will in principle have to be adapted for younger ages.

To date, the Levels of Personality Functioning Questionnaire for Adolescents (LoPF-Q 12-18) [21] is the only available self-report questionnaire to assess personality functioning according to the AMPD that was developed specifically for adolescents from 12 years up. The items of the LoPF-Q 12-18 have been carefully designed to take into account the developmental stage and life situation of adolescents [22]. It has been optimised for use in clinical practice, providing several descriptive subscales matching classical psychological concepts in addition to the total score and the four domain scores. This is supposed to inform differentiated diagnoses and therapy planning and to facilitate the upcoming fundamental changes in diagnostic guidelines for PD. First developed in German language, it has been translated and culturally adapted by expert teams for English [23], Spanish [24], Turkish [25,26] and Lithuanian [27].

Adaptations for Slovenian, Russian, French, Danish, Swahili (Tanzania), Hebrew, Chinese and Romanian are currently under development, showing that this instrument is supported by an international clinical and research community including low- and middle-income countries. The LoPF-Q 12-18 shows excellent scale reliability and accurately detects patients with personality disorders [21]. It can be requested for free for research purposes and is also available in electronic format at the project website (academic-tests.com).

### Test construction and psychometric properties of the LoPF-Q 12-18

The LoPF-Q 12-18 is a 97-item self-report measure for adolescents between 12 and 18 years (+/-2 years) to assess the dimensions of personality functioning: Identity, Self-direction, Empathy, and Intimacy. It is designed to enable a dimensional differentiation between healthy and impaired personality functioning to promote early detection of PD (criterion A). The construction was inspired by the AMPD [28] and the ICD-11 beta draft capturing the full scope of self- and interpersonal functioning. To operationalize the LoPF-Q 12-18, all descriptors of the four AMPD domains were carefully analyzed and enriched with available concepts from child and adolescent psychology with focus on clinical validity. This led to a detailed structure for operationalizing the domains of functioning (see S1 Table), building the basis for a deductive item formulation. The derived item pool was then revised in an empirically informed iterative process to make them appropriate for a self-rating instrument for adolescents. Accordingly, the four resulting primary scales identity, self-direction, empathy, and intimacy are composed of two subscales per scale. These subscales are reported in addition to the total score and scale scores to support detailed clinical decision making. Because they represent less abstract and more commonly shared concepts (like e.g. Purposefulness or Prosociality), they may be helpful to better understand a patient’s situation or to trace developments over time.

The process of test construction as well as psychometric properties have been described in detail in [22]. The main psychometric targets were clinical validity, good applicability for older and younger adolescents, and good scale reliabilities. The LoPF-Q 12-18 shows good scale reliability (Cronbach’s alpha of .96 for the total scale, .92, .94, .87, and .92 for the primary scales and between .76 and .96 for the subscales), good construct validity and substantial clinical validity. The LoPF-Q 12-18 total score distinguished between adolescents from the general population and n = 96 SCID-II diagnosed PD patients at a highly significant level and with a large effect size of 2.1 standard deviations [21].

As all four dimensions of personality functioning were designed to build upon the joint construct of PD severity, and since the AMPD defines a current PD as the presence of impairments in two or more of them, scales were expected and found to be highly intercorrelated (Pearson correlation coefficients ranged between .41 and .83). Exploratory factor analysis on item level supported a one-factor solution (i.e., strong first factor and a ratio of first to second factors’ eigenvalue of 5.1) speaking for a common factor of “personality pathology”. This is intended and in line with the goal of creating an assessment of the generalised severity of personality pathology. However, all four domains of functioning had been operationalized independently and in careful contrast to each other to make sure that each domain only covers one of the described aspects of PD-related impairments with minimum overlap. Each item had to show: sufficient item-total correlation as part of the assigned a) subscale, b) primary scale, and c) total scale, respecting an internal consistent structure on all scale levels, and a reasonable effect size for discriminating the school population and the PD patient sample as a sign of clinical validity. Factor analytic approaches were not used to empirically select the final item set. However, in an exploratory factor analysis on item level, a model with four factors accounted for 39.9% of the variance, and 72.2% of the items showed a loading > .30 on the factor that corresponded to the theoretically assumed domain. This was interpreted as preliminary evidence for the appropriateness of using the four domain scores [22]. However, with a Turkish translation of the LoPF-Q 12-18, a four-factor model did not show adequate fit in a CFA [26]. Therefore, the factor-analytical basis of the four domain scores has not yet been fully clarified.

### The Current Study

The first goal of this study was an in-depth investigation of the factorial structure of the LoPF-Q 12-18 items. Based on the preliminary analyses reported above, we expected that the LoPF-Q 12-18 is essentially unidimensional, in the sense that most of the reliable variance of the total score is due to a general factor. This is in line with research showing that different measures of PD severity capture a strong common factor and can therefore be scaled along a single latent continuum [29]. Nevertheless, previous research also indicates that specific factors might still play a role even when a strong first factor is present [30–33]. Hence, Goth et al., (23, p. 687) hypothesized that a bifactor structure might be suitable for the LoPF-Q 12-18, taking into account a strong general factor as well as four empirically distinguishable domains.

Our second goal for the current study was to achieve a considerably shorter version which maintains the structure of the questionnaire in terms of the four domains and the high clinical validity of the original long version. With 97 5-point likert scale items, the LOPF-Q 12-18 can be considered a somewhat long measure, at least for many research and clinical applications with a focus on fast and efficient screening. Length can, therefore, be considered a barrier for its usage. For instance, individuals with mental health problems often present in non-specialised settings like primary care, school psychologist offices, or emergency departments [34]. A shorter version would allow for administration of the instrument in a resource saving manner. This is important, as the LoPF-Q 12-18 might not be the only instrument that needs to be administered at a certain time. A short version can reduce burden for the patients and, additionally, it reduces resources required for the scoring of the questionnaire. Taken together, we expect that a short version will have a high impact on the practicability of the instrument.

## Materials and methods

### Participants and procedures

The current analyses were conducted on the same samples previously described in Goth et al. 2018 [22]. In short, a school sample of n = 351 students was assessed at three public schools. The BPFSC-11 (Borderline Personality Features Scale for Children, 11 Item Version; [35]) was used to screen for the PD related health status, n = 337 were below the Cut-Off ≥ 34 and was taken as healthy control group. The study was reviewed and approved by the ethics committee “Ethikkommission Beider Basel” which is now “Ethikkommission Nordwest-und Zentralschweiz”. Written informed consent has been obtained from all participants. A clinical sample of n = 415 patients was recruited at inpatient and outpatient units of six child and adolescent psychiatric hospitals in Basel, Innsbruck, Berlin, Mainz, Idar-Oberstein, and Heidelberg. Inclusion criteria were age of 12 to 20 years, sufficient language and cognitive skills, no autistic disorder, and no current psychotic episodes. Diagnoses were based on the results of the clinical interviews Structured Clinical Interview for DSM–IV Axis II (SCID–II; [36]), the Children’s Diagnostic Interview for Psychiatric Diseases (K-DIPS; [37]) and a classification conference. Patients with a PD diagnosis were assigned to the PD group independently from Axis I diagnoses. Of the total clinical sample, n = 96 patients (23.1%) met the DSM–IV criteria of one or more PDs (44.8% BPD). The total sample of n = 766 adolescents consists of 44.4% boys and 55.6% girls, the age range was 12-20 years (M = 15.5, SD = 1.9). For details, please see the description of the full study [22].

### Measures

The LoPF-Q 12-18 [21] has been described above. It contains 97 items to be answered on a 5-point scale ranging from 0 (no), 1 (more no), 2 (part/part), 3 (more yes) to 4 (yes). The resulting four scales Identity, Self-Direction, Empathy, and Intimacy are coded towards pathology and add up to a total score Personality Functioning, ranging from no impairment to severe impairment. For descriptive reasons, two subscales per scale are included, matching classical psychological concepts to facilitate interpretation. The test is available on the self-publishing project website (academic-tests.com).

### Investigation of the latent structure

For the investigation of the latent structure of the LoPF-Q 12-18, confirmatory factor analyses on item level were used. Scale reliabilities were evaluated using McDonald’s Omega. The analyses were conducted with the software ‘R’ [38] and the package ‘lavaan’ [39]. Fig 1 illustrates models representing different factorial assumptions that were tested in order to compare their fit.

**Fig 1:**
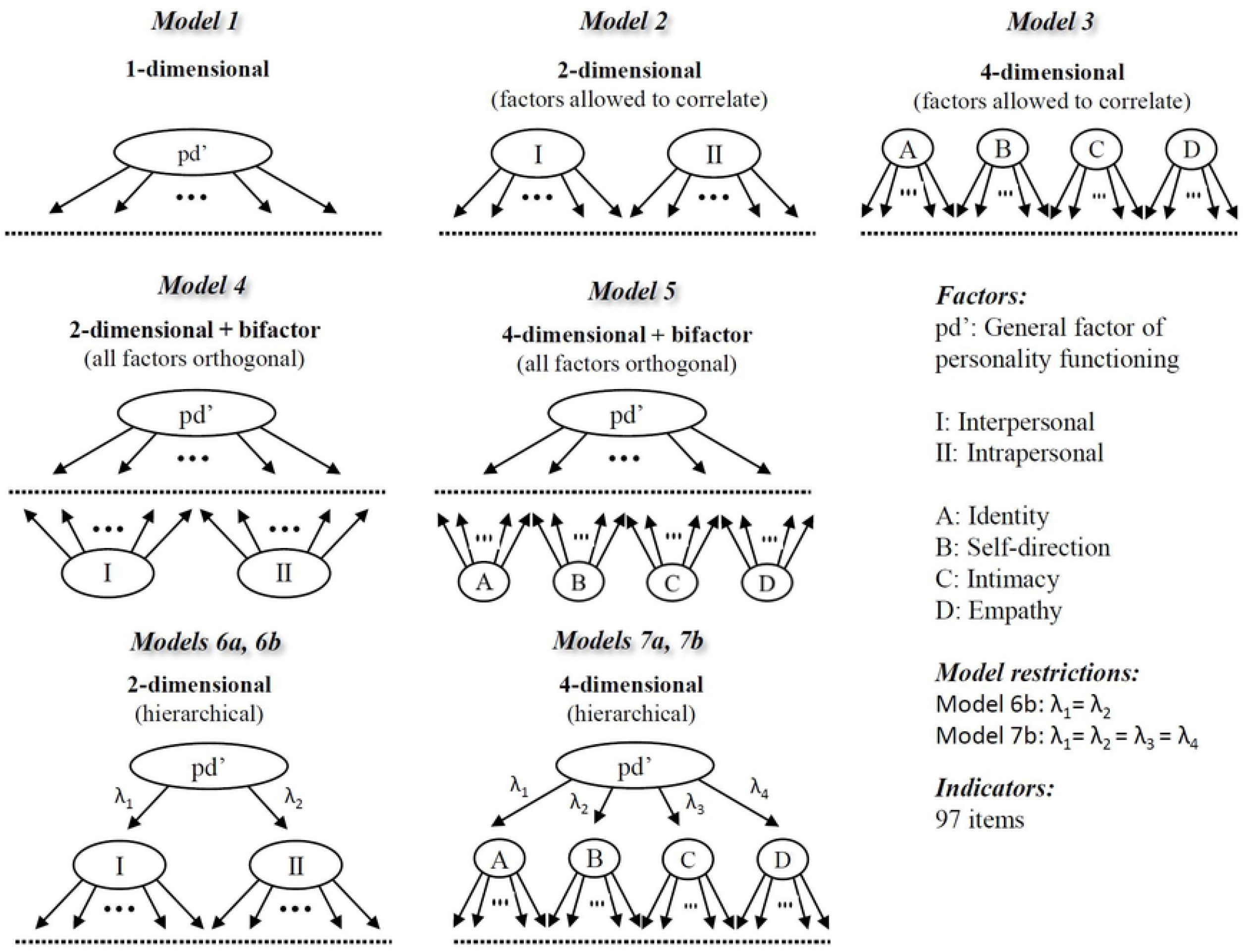
Different configural assumptions tested for the long version

The following fit indices are reported: Comparative Fit Index (CFI), Root Mean Squared Error of Approximation (RMSEA) and Standardized Root Mean Square Residual (SRMR) [40]. The following combination of indices was used as the cut-off to determine acceptable models: CFI >. 90 acceptable and CFI >.95 good, RMSEA <.05, SRMR <.08 [41,42]. The model fit of the short version created by ACO method (see below) was investigated using the same criteria. Scale reliabilities were estimated using the package ‘semTools’ [43]. We report an ordinal version of coefficient alpha according to Zumbo et. al [44], as well as omega hierarchical according to McDonald [45]. Note that in bifactor models, omega hierarchical corresponds to OmegaH for the total score and to OmegaHS for the subscale scores [46].

### Ant Colony Optimization to create a short version

For small item pools it can be an option to iterate through all possible item combinations or to apply simple iterative methods, e.g. a Stepwise Confirmatory Factor Analysis Approach (SCOFA) in order to find a well-suited combination of items that can be used as a short version of a test [47]. Considering that the LoPF-Q 12-18 consists of 97 items, the number of possible item combinations reaches a level where this is no longer possible. To illustrate that such an attempt is impossible, we calculated the number of possible combinations based on “n over k”, as suggested previously [48]. The S2 Table illustrates the estimated computation time, memory, and amount of energy that would be required to execute these iterations. Results show that the algorithm would need to run for billions of years while using multiple times the global estimated yearly energy consumption. Consequently, it is inevitable to run an optimization algorithm which approximates a close-to-optimal short version of the questionnaire in a shorter period of time.

The Ant Colony Optimization (ACO) meta-heuristic [49] was used to select a set of items for the short version. For our use case, this algorithm ran for less than 5 hours CPU time (see S2 Table). The ACO consists of virtual ‘ants’ who explore the selection of sets of items and attribute a ‘pheromone’ level to the items of the selection according to a statistical criterion (defined below). Items with a higher pheromone level have a higher probability to be re-selected in future item sets to be explored. The pheromone level fades over time (“evaporation”). The ACO was started with 30 ants, and the algorithm stopped after 20 runs without improvement. This method has already proven useful in the construction of the 34-item “Personality Inventory for DSM-5, Brief Form Plus” (PID5BF+) [50].

Our goal was to generate a short version of the LoPF-Q 12-18 with a total of 20 items, including 5 items for each domain (Identity, Self-direction, Intimacy, Empathy). For this purpose, the ACO method was set up to select a subset of 5 items from each domain of the original version. The criterion to calculate the pheromone used by the ants was a combination of model fit, reliabilities of the domain scales and clinical validity i.e. the capacity to discriminate between patients with personality disorder (n = 96) and students without signs of personality disorder according to the BPFSC-11 (n = 337). Model fit was based on a confirmatory factor analysis with 4 first order factors to represent the domains and a secondary higher order factor to represent generalised severity. The loadings of the domains on the higher order factor were constrained to be equal, thereby ensuring a balanced interpretation of the general severity continuum. Pheromones were calculated based on logistic transformations (φ) of fit measures (CFI, RMSEA), measures of reliability (McDonald’s Omega and minimum factor loading) and criterion validity (adjusted R2). The ultimately optimised (i.e. maximised) pheromone was based on the sum of all three φ-values. Please refer to S3 Materials for the formulas used for calculating pheromone levels.

Finally, to show the advantage of the applied ACO-algorithm over the iterative approach, we compared reliability, CFA model fit as well as criterion validity with 100,000 random combinations of items. Due to the high computational load, the calculation of these 100,000 models was performed at the sciCORE (http://scicore.unibas.ch/), the scientific computing centre of the University of Basel. All analyses have been conducted using R (> version 4.0.1), as well as the R packages ‘lavaan’ [39] and ‘semTools’ [43] for Confirmatory Factor Analyses. Receiver operating characteristic (ROC) analysis was used to determine the clinical utility of the LoPF-Q short version and to derive empirical cut-off scores for defining clinically relevant thresholds.

## Results

### Investigation of the factorial structure of the original (97-item) version

Table 1 shows the parameters of different confirmatory factor analyses (CFA) testing different factorial assumptions.

**Table 1:** Confirmatory Factor Analyses (CFA) testing different factorial assumptions (long version)

| Model (id) | Factors | par | $\chi^2$ | CFI | RMSEA | SRMR |
| --- | --- | --- | --- | --- | --- | --- |
| 1-dim (1) | 1 | 485 | 12341.2 | 0.851 | 0.057 | 0.090 |
| 2-dim (2) | 2 | 486 | 11584.6 | 0.865 | 0.054 | 0.087 |
| 4-dim (3) | 4 | 491 | 11019.2 | 0.876 | 0.052 | 0.084 |
| 2+bifactor (4) | 2 | 582 | 9286.3 | 0.907 | 0.046 | 0.070* |
| 4+bifactor (5) | 4 | 582 | 9099.5* | 0.911* | 0.045* | 0.072 |
| 2-dim hierarchical (6a)<sup>i</sup> | (2) | (487) | (21882.8) | (0.667) | (0.085) | (0.087) |
| 2-dim hierarchical (6b) | 2 | 486 | 11584.6 | 0.865 | 0.054 | 0.087 |
| 4-dim hierarchical (7a) | 4 | 489 | 11089.0 | 0.875 | 0.052 | 0.085 |
| 4-dim hierarchical (7b) | 4 | 486 | 11397.4 | 0.869 | 0.054 | 0.086 |

Note. *npar*: number of estimated parameters; *df*: degrees of freedom; *CFI*: Comparative Fit Index; *RMSEA*: Root Mean Squared Error of Approximation; *SRMR*: Standard Root Mean Residual. Best fit indices are highlighted with asterisks. <sup>i</sup> For model 6a standard errors could not be computed and the information matrix could not be inverted. This may be a symptom of a non-identifiable model. Therefore, the parameters of this model are shown in parenthesis.

All factorial assumptions are related to the basic personality functioning concept, highlighting either the joint construct of PD severity, the two areas of Self-related and Interpersonal functioning, or the four domains Identity, Self-Direction, Empathy, and Intimacy according to the AMPD. Overall, the bifactor models performed slightly better than all other correlated or hierarchical factor models. A four-dimensional bifactor was the only model to show acceptable fit based on all three fit measures, RMSEA (<.05) and SRMR (<.08) and CFI (>.90). The two best fitting bifactor models (“two-dimensional bifactor” and “four-dimensional bifactor”) showed very similar fit indices with only a subtle difference on RMSEA. Table 2 summarizes the model-based scale reliabilities. Based on the best fitting model (model 5), ordinal alpha was excellent for the general factor as well as all four domains (>.90). However, while OmegaH showed excellent reliability of the total score (.94), OmegaHS was substantially lower for the four domains scales (.07, .11, .50, .20) (see S4 Tables – sheet 1 for additional details). Factor loadings of model 5 indicated that several items from the domain of empathy did not substantially (> .30) load on the general factor, and several items from the domain of identity had even negative loadings on the respective specific factor (see S4 Tables – sheet 2). S4 Tables – sheet 3 shows the factor intercorrelations for the bi-factor models.

**Table 2:**
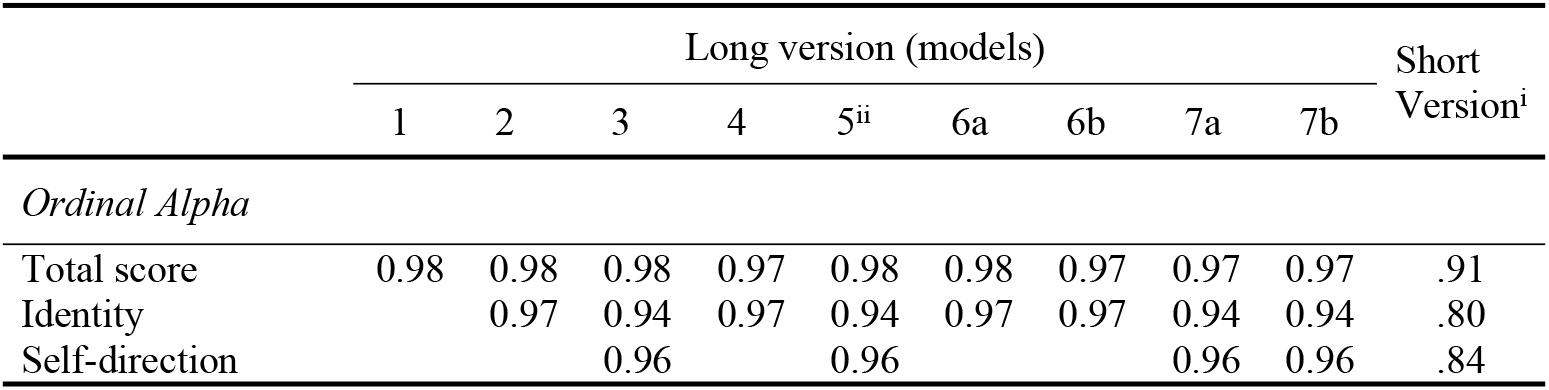
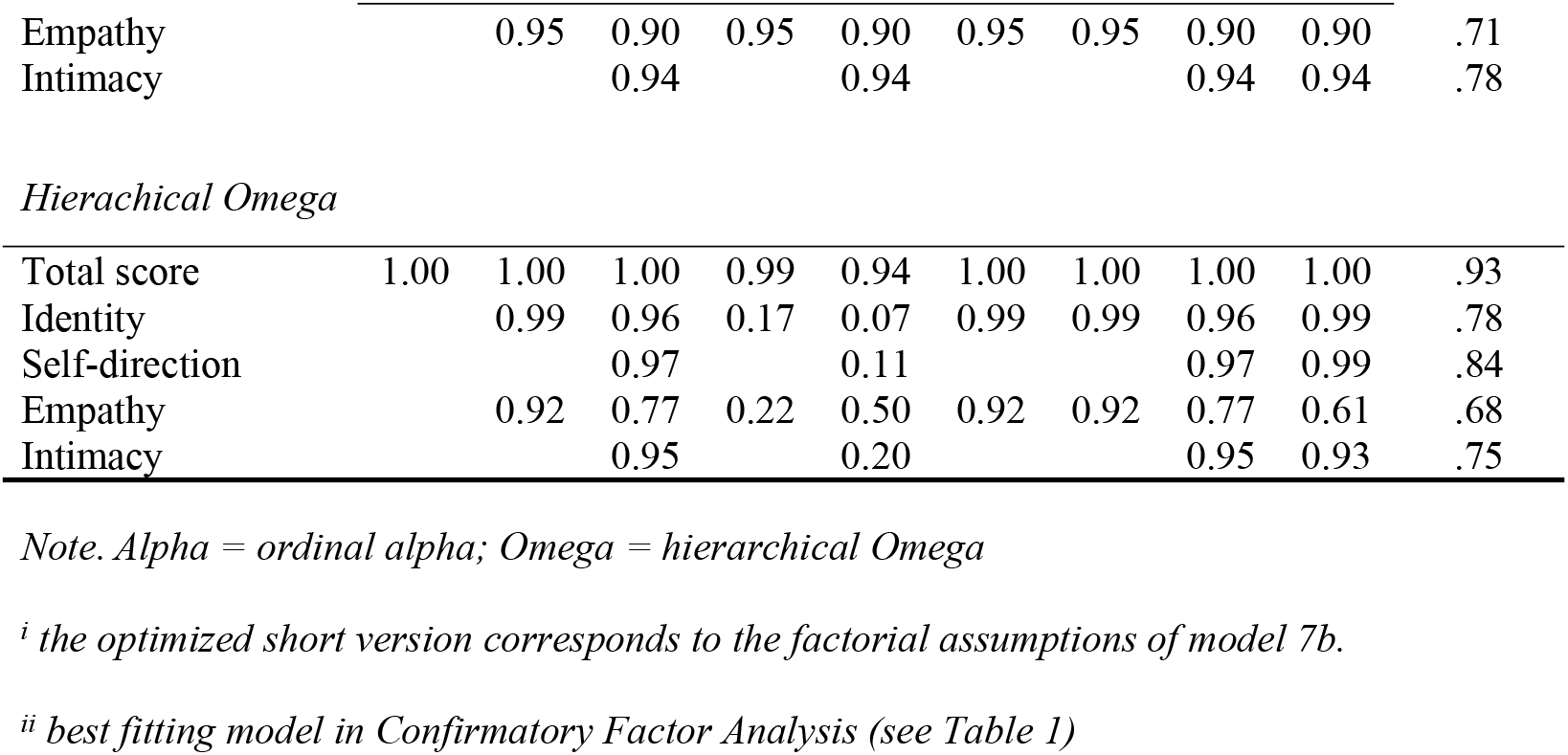
Factor reliabilities for the long and short version

Taken together, this suggests that although model 5 has the best fit, it is not a very satisfying representation of the structure of the LoPF-Q 12-18 [51]. In consequence, we have chosen a hierarchical model with four lower-order factors (i.e., model 7b) for developing the short version.

### Creating a short version

As intended, a 20-item version with 5 items for each of the domain scales was obtained. Fit indices presented in Table 3 show that the optimised short version had a very good fit on all fit indices (CFI = .980, RMSEA = .046, SRMR = .038).

**Table 3:**
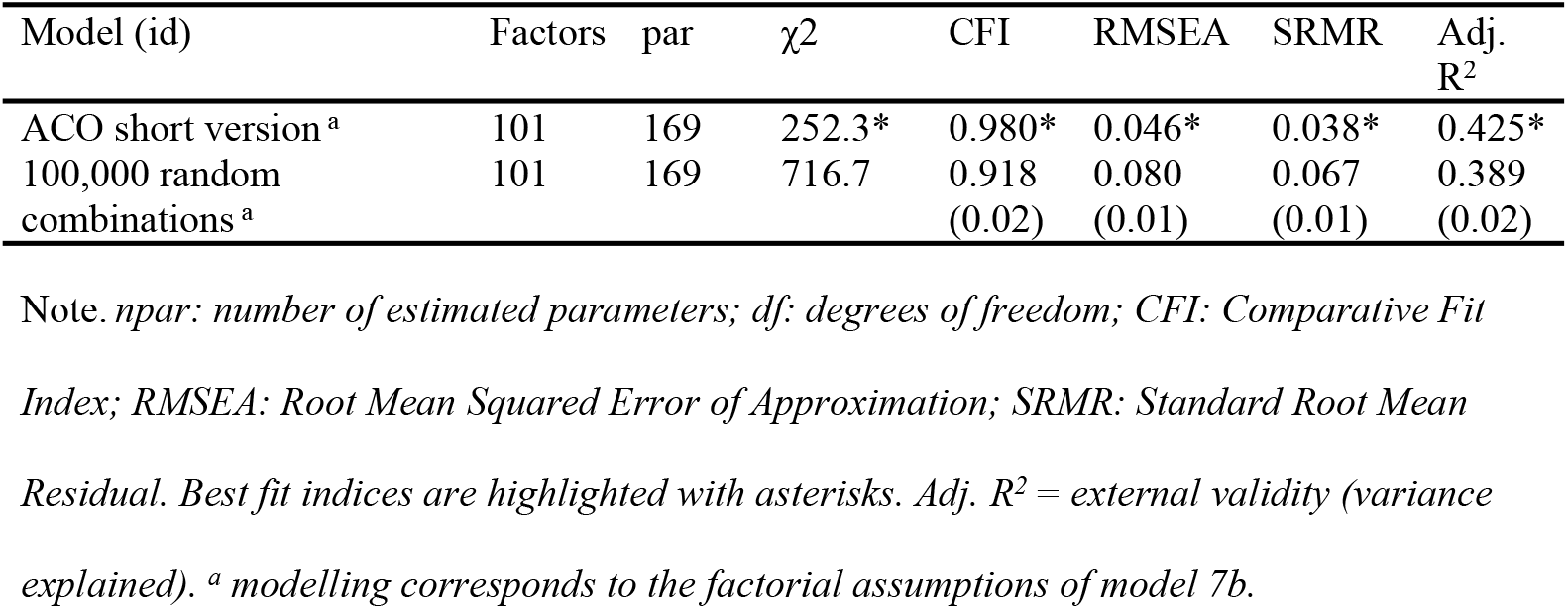
Model fit indices and external validity for the Ant Colony Optimised Short version compared to 100,000 random combinations of items

Due to the smaller number of items, ordinal alpha was slightly lower in comparison to the long version (.91 total scale, .71 - .84 domains). Omega hierarchical of the total score was similar (.93). Factor loadings of the optimised short version are depicted in Fig 2.

**Fig 2:**
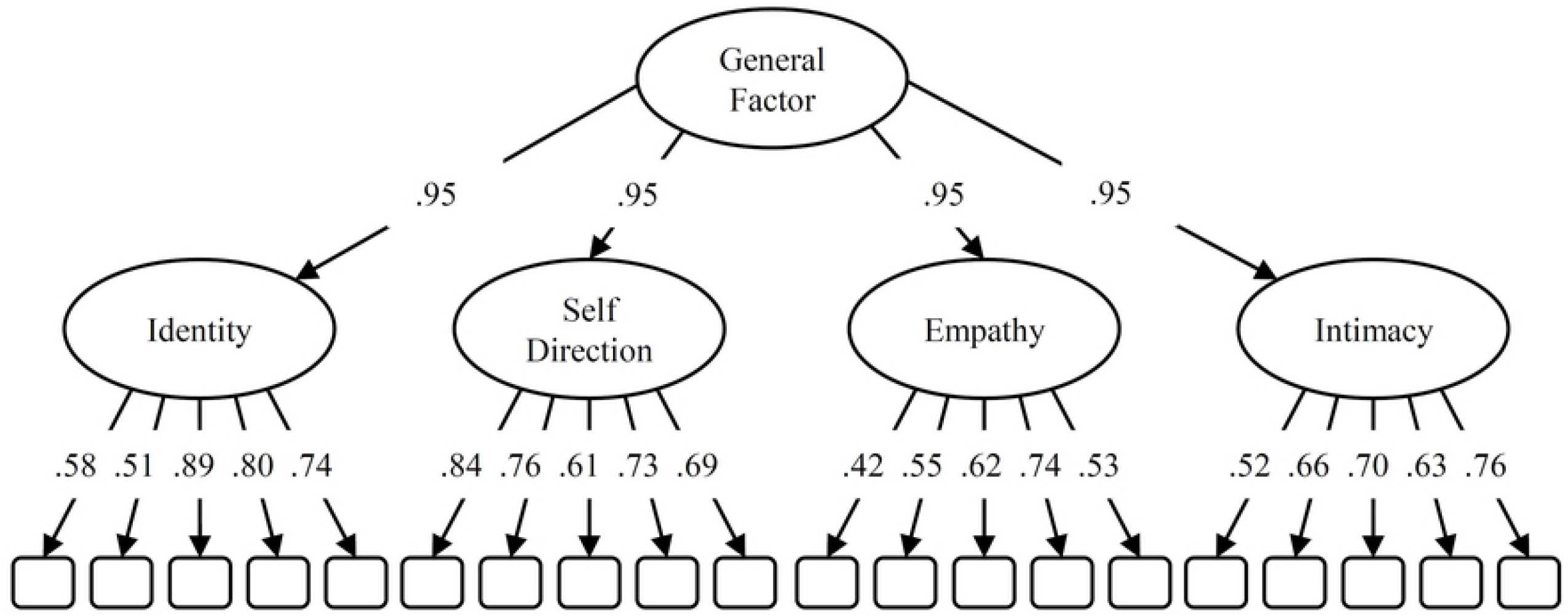
Factor loadings of short version. Optimized model using ant colony optimization to develop a short version to identify personality disorders in adolescence. The configuration corresponds to model 7b in Fig 1

The final short version was optimized in a way that the lower-order factor loadings were kept constant across all four domains. This procedure yielded very high and equidistant factor loadings (.95) on the general factor.

For comparison, Table 2 additionally shows the average model fits (and standard deviations) for 100,000 randomly selected item combinations testing the same hierarchical factor model. Fig 3 visually compares the 20-item solution that was generated with the ACO with 100’000 random combinations of items regarding external validity and model fit. Compared to the random combinations, the combination of model fit and external validity (the ability to differentiate between healthy controls and PD patients) of the short ACO version are excellent with an adjusted R square of .425 (i.e. 42,5% explained variance).

**Fig 3:**
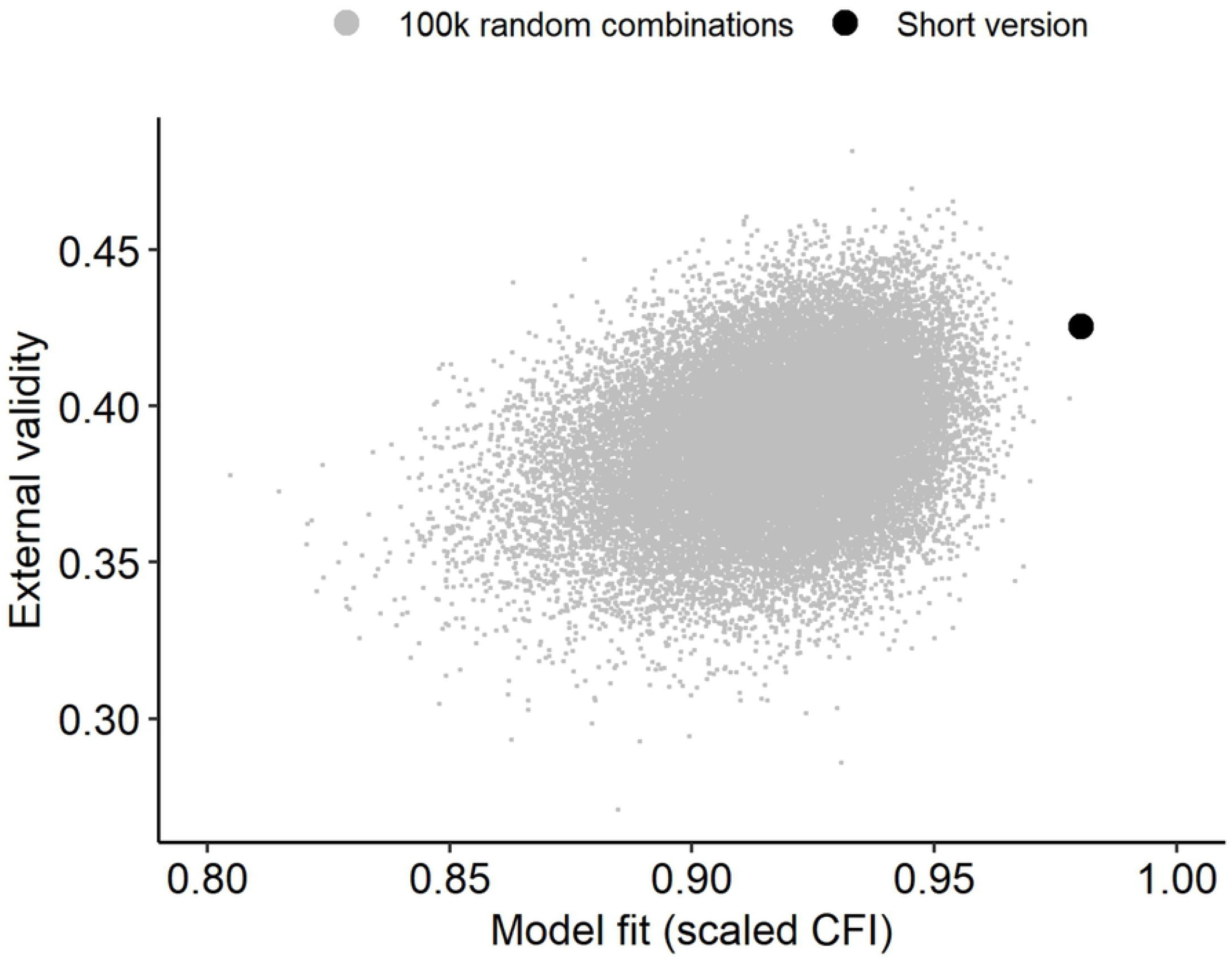
Fit and external validity of short version. Model fit and external validity of the optimized short version in comparison to 100,000 random item combinations.

Expressed with the more traditional effect size Cohens d, the LoPF-Q Short total score differs between the PD patients and the healthy controls with d = 3.1 standard deviations. ROC analysis showed an area under the curve (AUC) of .92 (p < .001; 95 % confidence interval .89-.95). A preliminary cut-off score for the LoPF-Q Short total score was defined to be ≥ 36 using Youden index, corresponding to a T-score of 74. Specificity for detecting patients with personality disorder compared to healthy controls was 87.5% and sensitivity was 80.2%. Reliability coefficients for the short version can be seen in Table 2.

## Discussion

The current study had two aims: First, to investigate the latent structure of the LoPF-Q 12-18 original 97-item version. We assumed that a bifactor model with a general factor – representing a joint construct of PD severity – with four specific factors matching the four domains of functioning according the AMPD (DSM-5) would perform well. The second goal was the construction of an optimised short version to meet the needs for an efficient screening instrument for PD in adolescents.

### Factorial structure of LoPF-Q 12-18 long version

As hypothesised, the nominally best fitting models were bifactor models when compared to correlated factor or hierarchical factor models (see Fig 1). The model fit of both bifactor models (including either two broad dimensions or four narrow domains) was acceptable when considering all evaluated criteria. However, other aspects besides model fit should be considered when interpreting bifactor models [51] due i.a. to their less restrictive nature which results in a higher overall chance of good fit, even when using random data. In addition to an acceptable fit, the items should show substantial loadings on the general factor, and the specific scales should have sufficient reliability after controlling for variance of the general factor (i.e., Omega HS). In both these respects the estimates of the bi-factor models were lacking. Conclusively, bi-factor models did not satisfactorily represent the structure of the questionnaire despite the acceptable fit. Nevertheless, the following important conclusions can be drawn from the performed analyses.

First, as expected, the item level data collected with the LoPF-Q 12-18 contain a very strong general factor. This can be seen, for example, in the fact that model fit was only moderately improved by extracting more than one factor, that the four domains in model 3 were very highly correlated (S4 Tables – sheet 3), and that the reliable variance in the total score in model 5 (i.e., Omega H) was almost entirely attributable to the general factor (S4 Tables – sheet 1). This support for a general factor of personality functioning is very much in line with the usage of the LoPF-Q 12-18 in the framework of diagnostic procedures of both the AMPD (DSM-5) and the ICD-11. In both diagnostic models, personality functioning is seen as an overarching construct important to establish a PD diagnosis and to judge its severity. Importantly, according to [52], the general factor of personality pathology has been primarily described in adult populations. The current study might be one of the first to describe this general factor in a sample of younger patients.

Second, the four domain subscales, with the possible exception of empathy, contain hardly any reliable variance beyond general severity. In other words: Although the four domain scores were reliable in their own right (i.e., ordinal alpha > .90), their very high correlation in the underlying sample makes it seem unlikely that distinctive and clinically interpretable profiles will emerge in individual cases. This contrasts with PD criteria from DSM-IV [32,53] or items of the Inventory of Personality Organization [31], which tend to warrant scoring of subscales in adult samples. At least on a group level, [22] found first evidence of distinctive profiles, for example, the empathy scale was severely impaired only in patients with narcissistic and antisocial PD, whereas the identity scale was particularly impaired in patients with Borderline or anxious-avoidant PD. The specific clinical variation of the empathy scale may be an explanation for why only this one showed an independent variance beyond the general factor. In sum, whether the use of each of the four domain scores is clinically meaningful needs to be investigated in clinical trials with different types of PD patients and optimally with different therapeutic approaches in a longitudinal design.

The debate on the meaningfulness of the domain scales is important as mental health care workers tend to find the primary scales and subscales of the LoPF-Q 12-18 useful for the interpretation of the assessments regarding clinical decision making and therapy planning. This is comprehensible as the less abstract denomination of the subscales appear to be closer to commonly shared concepts and can be used to find a shared language with the patients and their families. According to the authors [22] the LoPF-Q 12 -18 has been primarily developed to meet the needs of clinical practitioners and to cover a wide range of symptoms related to the four domains, because often specific aspects of functioning (identity pathology, problems with self-regulation or problems with social interaction etc.) are the primary target for psychotherapy. This discrepancy between the authors’ experiences and intentions and our current findings cannot be conclusively clarified. The currently investigated sample consisted mainly of subjects without signs of PD (351 from schools and 319 patients without PD vs 96 patients with PD). The general factor might turn out being less pronounced and the domain scales more independent from each other when investigating clinical samples of PD patients [54]. Similarly, Watts et al. found that the inclusion of undiagnosed individuals causes more positive correlations in psychopathological data, leading to a stronger *p*-factor [55]. For a further optimization of the structure in a short form of the LoPF-Q 12-18, it seemed reasonable to keep the four domains in terms of content validity, but to put the focus of the optimization on the general factor.

### LoPF-Q Screener (20-item version)

The short version was optimised for clinical validity and internal consistency accounting for a structure with four first order factors to represent the personality functioning domains and a secondary higher order factor to represent the general personality functioning denoting PD severity. Thanks to the optimisation, the short version performed excellently regarding both external validity and internal consistency. The optimisation was done with the ACO heuristic which had already proven useful in previous studies for creating short personality assessments [31,50] and performed very well in the current study (see Fig 3). The derived short version “LoPF-Q Screener” contains 20 items and preserves the four scales Identity, Self-direction, Empathy and Intimacy as well as the total scale Personality Functioning. It showed an excellent model fit concerning all parameters and good scale reliabilities. Most importantly, it showed excellent clinical validity, with the total scale differentiating significantly and with an effect size of 3.1 standard deviations between PD patients and healthy adolescents.

The LoPF-Q Screener can be used in contexts where employing the longer version is not feasible or inconvenient. This flexibility cannot be overestimated in the presence of a general global mental health gap [56] in adolescents and a specific gap regarding personality disorders in youth [6,7]. Tools that can help address these gaps are required, and while diagnostic tools cannot solve this issue alone, they are one of the cornerstones to advance research and interventions. The results on psychometric properties of this short version are still preliminary and need to be verified with test data that were not used for its construction. The data ideally needs to be collected with this short version in order to validate it, since using a subsample of items of data collected with the long version might potentially introduce bias (e.g. memory effects, effects of the sequential order of items, attention span of the subject etc.). Finally, the question arises whether an even shorter version wouldn’t be better in terms of practicality of the assessment and, thus, versatility in clinical contexts. However, an even shorter version may come at the expense of inferior measurement precision and diagnostic validity, both of which are highly relevant for clinical usage [57]. The 20-item version of the LoPF Q 12-18 is likely to present a solid compromise between psychometric precision and practicality.

### Research recommendations

Research on the usefulness of the levels of personality functioning model for clinical decision making such as selection of appropriate treatment and treatment customisation is needed. The long and short version need to be compared in future studies regarding their usability and user experience of the different stakeholders. For instance, do users benefit from the more comprehensive data collection of the long version or are they looking for more efficient tools? A further question is the preparation of a pathway towards shorter versions for different cultural settings. The authors of the LoPF-Q 12-18 pursue a strategy in which they emphasise the importance of the same set of items for all cultural settings and actively support the development of cultural adaptations and networking among interested colleagues. A shared set of items across culturally adapted versions is necessary because it facilitates scientific exchange and management of the different versions and enables joint data analyses in cross-cultural settings.

This possibility is particularly important because the development of PD in early adolescence is an under-researched area and data pooling is key. In addition, LoPF-Q versions for informant report and for even younger age groups (from 6 years up) are under development, and the seamless and clear transferability of the assessed scales in all cultural adaptations is crucial, especially for longitudinal studies. Future research will show whether the optimised short version LoPF-Q Screener will provide measurement invariance across different cultural settings and translations.

The current study highlights the usefulness of a more detailed and more time-efficient assessment of personality functioning in adolescence. Whereas there is no doubt about a common core, i.e. a general latent construct, there is somewhat mixed evidence regarding the usefulness of the lower-order domains (identity, self-direction, empathy, intimacy). Earlier research on alcohol use disorders has shown that determining the factor structure in a sample including individuals with no clinical symptoms may have a debilitating impact on the discrimination of sub-factors [54]. Future research on the LoPF-Q 12-18 and the introduced LoPF-Q Screener short version might provide more comprehensive insights by comparing the factor structure between clinical and non-clinical samples.

## Data Availability

Data cannot be shared publicly because the consent forms did not contain this possibliy. Data are available from the last author KG. KG can be contacted via this form: https://academic-tests.com/contact/

